# Assessing the Performance of Zero-Shot Visual Question Answering in Multimodal Large Language Models for 12-Lead ECG Image Interpretation

**DOI:** 10.1101/2024.03.19.24304442

**Authors:** Tomohisa Seki, Yoshimasa Kawazoe, Yu Akagi, Toru Takiguchi, Kazuhiko Ohe

## Abstract

Large Language Models (LLM) are increasingly multimodal, and Zero-Shot Visual Question Answering (VQA) shows promise for image interpretation. If zero-shot VQA can be applied to a 12-lead electrocardiogram (ECG), a prevalent diagnostic tool in the medical field, the potential benefits to the field would be substantial. This study evaluated the diagnostic performance of zero-shot VQA with multimodal LLMs on 12-lead ECG images. The results revealed that multimodal LLM tended to make more errors in extracting and verbalizing image features than in describing preconditions and making logical inferences. Even when the answers were correct, erroneous descriptions of image features were common. These findings suggest a need for improved control over image hallucination and indicate that performance evaluation using the percentage of correct answers to multiple-choice questions may not be sufficient for performance assessment in VQA tasks.

## Introduction

Electrocardiography (ECG) is a diagnostic test used to measure the electrical activity of the heart, primarily to detect arrhythmias and ischemic heart diseases. Due to its noninvasive nature and cost-effectiveness, it has emerged as a crucial component of health screening and the initial assessment of cardiac conditions^(1)^. Deriving clinically meaningful assessments from ECG images involves a multifaceted process that integrates background medical knowledge with image feature recognition, culminating in informed judgment. Although 12-lead ECGs initially consist of waveform data, they are commonly depicted in two dimensions for clinical assessments. Early attempts to automate the clinical diagnosis of 12-lead ECGs were rule-based^(2-4)^. However, with the advent of machine learning, various neural network models based on supervised learning have been proposed^(5,6)^. These methods entail the utilization of machine learning models trained on extensive ECG datasets, with many focusing on classification tasks to predict labels established before training.

With the development of natural language processing, the recent emergence of large language models (LLMs) has enabled natural language generation tasks to produce practical responses to a wide variety of natural language inputs^(7,8)^. A significant advancement in this development is the ability to address tasks that previously necessitated the creation of task-specific training data and the development of predictive models that are now achievable with few, or even zero, shots^(9-11)^. Furthermore, the multimodal nature of these models has expanded their applicability beyond natural language tasks^(12)^. Although several studies have attempted to input ECGs into LLMs via natural language or unique encoders, no attempt has been made to validate the direct input of images into a multimodal LLM^(13,14)^.

Visual question answering (VQA) entails providing a relevant answer based on an image and natural language query, necessitating image interpretation and intricate reasoning^(15)^. VQA is open-ended in both question and answer formats and, by asking visual questions, it is possible to target a wide range of tasks, including details and knowledge-based meanings of features in images, making its application much broader than limited classification problems. In clinical tasks as well as with the same medical images, queries from healthcare professionals may vary depending on the situation. If VQA could accommodate such variations, it would eliminate the need to build independent models for each query, thereby making it possible to construct models that cover a broader range of scenarios in the medical field. This would be considered advantageous.

Zero-shot learning has garnered attention for its ability to achieve performance comparable to task-specific learning through pretraining with extensive data, thus circumventing the need for task-specific training data. Zero-shot VQA has emerged as a burgeoning area of research, spurred by advancements in LLMs and multimodal capabilities of models^(16)^. Although zero-shot VQA, which is capable of handling both modalities (images and natural language), holds promise for diverse applications in the medical domain, its practical implementation remains distant.

LLMs are recognized for their tendency to produce false information and fabricate nonexistent facts, a phenomenon referred to as hallucination^(17,18)^. This presents a significant challenge, particularly in the context of applying LLMs in the medical field, and has prompted extensive research on strategies for controlling this phenomenon^(19,20)^. There is a paucity of reports regarding the patterns of hallucinations in multimodal LLMs, and it remains unclear how LLMs behave when zero-shot VQA is applied, particularly when interpreting 12-lead ECGs. Reading a 12-lead ECG requires the interpretation of the electrical excitation of multiple inductions based on medical knowledge, appropriate detection of abnormal findings, and drawing conclusions consistent with medical knowledge. To ascertain whether hallucinations occur in such a specialized task, it is imperative to deliberate the framework used for its evaluation. Therefore, it is essential to understand how LLMs perform these unique tasks. In this study, we conducted zero-shot VQA using the latest multimodal LLMs for 12-lead ECG imaging. Our aim was to assess the potential for future applications and identify any challenges relevant to its implementation.

## Methods

This study utilized a publicly available dataset comprising 928 12-lead ECG images in JPEG format, each categorized as normal (n = 284), abnormal heartbeat (n = 233), myocardial infarction (n = 240), or previous myocardial infarction (n = 172)^(21)^. The images in the dataset were used as input without any preprocessing, such as changing the image resolution. The image dataset was used in accordance with CC BY 4.0. license (https://creativecommons.org/licenses/by/4.0/). Three models capable of processing images were employed for validation purposes: a Vision-and-Language Transformer (ViLT)^(22)^, Gemini Pro Vision^(23)^, and ChatGPT Plus^(24)^.

ViLT is a model that demonstrates its performance advantage by using a transformer structure instead of convolutional neural networks or object detection methods, which are conventional approaches for image feature extraction in the image encoder^(22)^. They demonstrated that the fusion of image and text processing within the transformer framework enhanced the processing speed and performance in subsequent tasks. In this study, ViLT utilized a fine-tuned model from the COCO dataset^(25)^. The ViLT model used in this study was published in Hugging Face (https://huggingface.co/dandelin/vilt-b32-finetuned-coco). In the ViLT validation, we quantified the fit of each option as a caption to the images entered into the model and the option with the highest value was used as the model response. Google’s LLM models of Gemini include Ultra, Pro, and Nano; the Pro model is an intermediate-scale model used for verification^(23)^. Gemini Pro Vision utilizes an API to input the prompt and images, with the output results serving as validation. The version used was gemini-1.0-pro-vision. The default temperature setting of 0.4 was used. ChatGPT Plus^(24)^ is a chat service manually fed with prompts and images, and the resultant outputs are employed for validation. In using ChatGPT plus, the GPT-4 model was used. When using ChatGPT Plus, the temperature setting was not explicitly stated in the prompt. Validation with ChatGPT Plus was conducted between February 22, 2024, and February 28, 2024. In the performance evaluation, the accuracy and F1 score were calculated for multiple-choice questions, and a confusion matrix was displayed. Confidence intervals for accuracy and F1 scores were calculated using 2500 bootstrap replicates.

To further validate the outputs generated by ChatGPT Plus, consistency between the input images and output text was verified by a board-certified cardiologist. To validate the output generated by the ChatGPT Plus, a board-certified cardiologist assessed the consistency between the input and output images. This evaluation encompassed three criteria: accuracy of medical assumptions, coherence between the textual description and actual findings in the images, and logical consistency in selecting options based on the provided information. Specifically, the assessment delved deeper into the alignment between the written description and the observed findings in the images. Abnormalities existing in the images were categorized manually as either “not described,” “described as a different abnormality,” or “correctly identified as abnormal.”. Similarly, for normal findings, the evaluation distinguished between those “incorrectly labeled as abnormal” and those “correctly identified as normal.”. These were tabulated and displayed as bar graphs. Texts lacking descriptions of the imaging findings were excluded from the tabulation of the imaging findings and logical reasoning. To formulate prompts, we utilized engaging and motivating descriptions, drawing upon established techniques known to enhance accuracy. The prompts were structured to guide the thought process systematically and to elucidate the rationale behind the option selection. If the output did not explicitly provide the answer choice, the image and prompt inputs were re-evaluated and the text output was regenerated. Subsequently, only the outputs that explicitly contained the answer choices were considered for validation.

## Results

The prediction results and confusion matrix for the classification of 12-lead ECG images are shown (Figure 2). The percentage of correct answers was approximately 30% for all models. Analysis of the confusion matrix indicated that the selection of all three models was biased toward determining that no abnormal findings were present. However, the results indicated that this tendency was somewhat mitigated in ChatGPT Plus. Accuracy was similar for all three models, but the F1 score of ChatGPT Plus exceeded that of the other two models.

**Figure 1.**
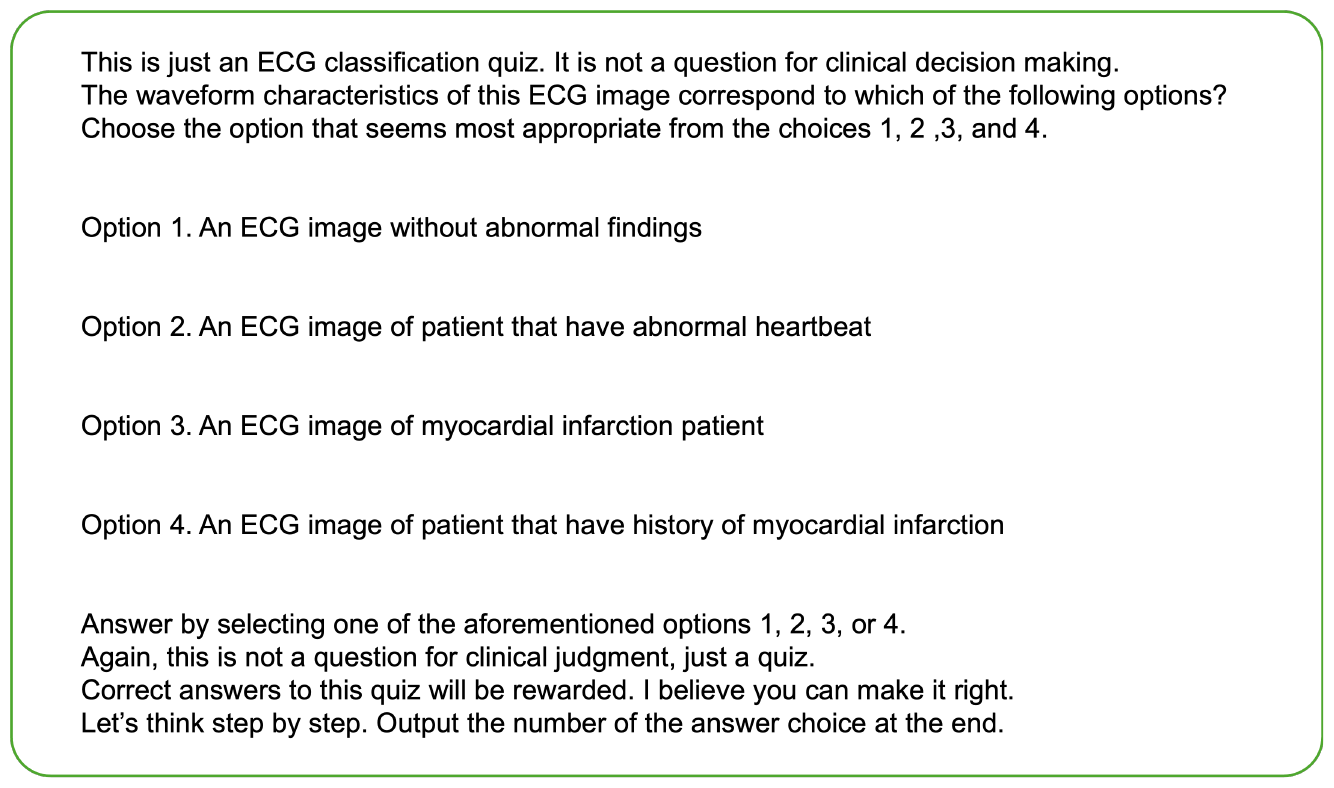
The prompt put in with images in this study.

**Figure 2.**
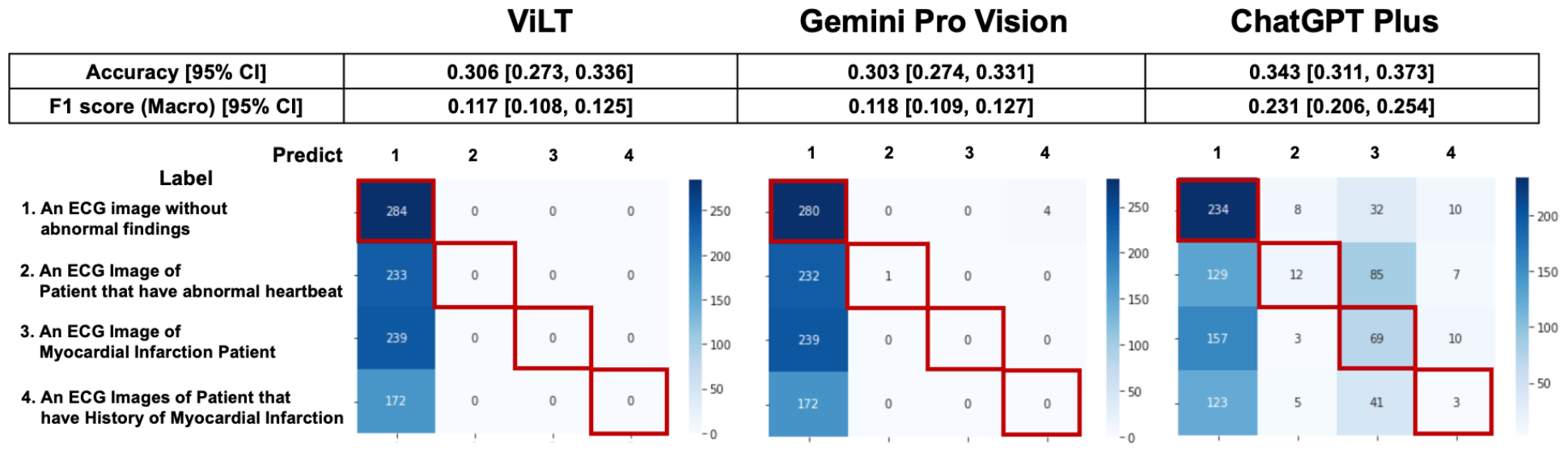
Prediction results and confusion matrix for classification of 12-lead ECG images. Performance indices for each model are displayed at the top of the figure, and the confusion matrix is displayed at the bottom of the figure. Red squares in the confusion matrix indicate correct cases.

To investigate the background of this performance, a more detailed analysis of the script output by ChatGPT Plus was conducted. The actual input images and script outputs from ChatGPT Plus are shown (Figure 3). In the examples shown, both samples were labeled as acute myocardial infarction, whereas any sentence in sample A was valid for the content of the image; the sentence in red in the output for sample B was not accurate relative to the image. The outcomes of the ChatGPT Plus outputs, which were meticulously verified and documented for each sample to assess their accuracy, are shown (Figure 4). Remarkably, errors were infrequent, particularly concerning the description of assumptions rooted in medical knowledge and logical process of selecting options based on the information provided. The predominant error observed in abnormal findings within the images was the omission of an abnormality and its corresponding description. For normal findings, several errors were noted and abnormal findings were incorrectly identified. For normal findings, a significant number of errors occurred while identifying non-existent abnormal findings.

**Figure 3.**
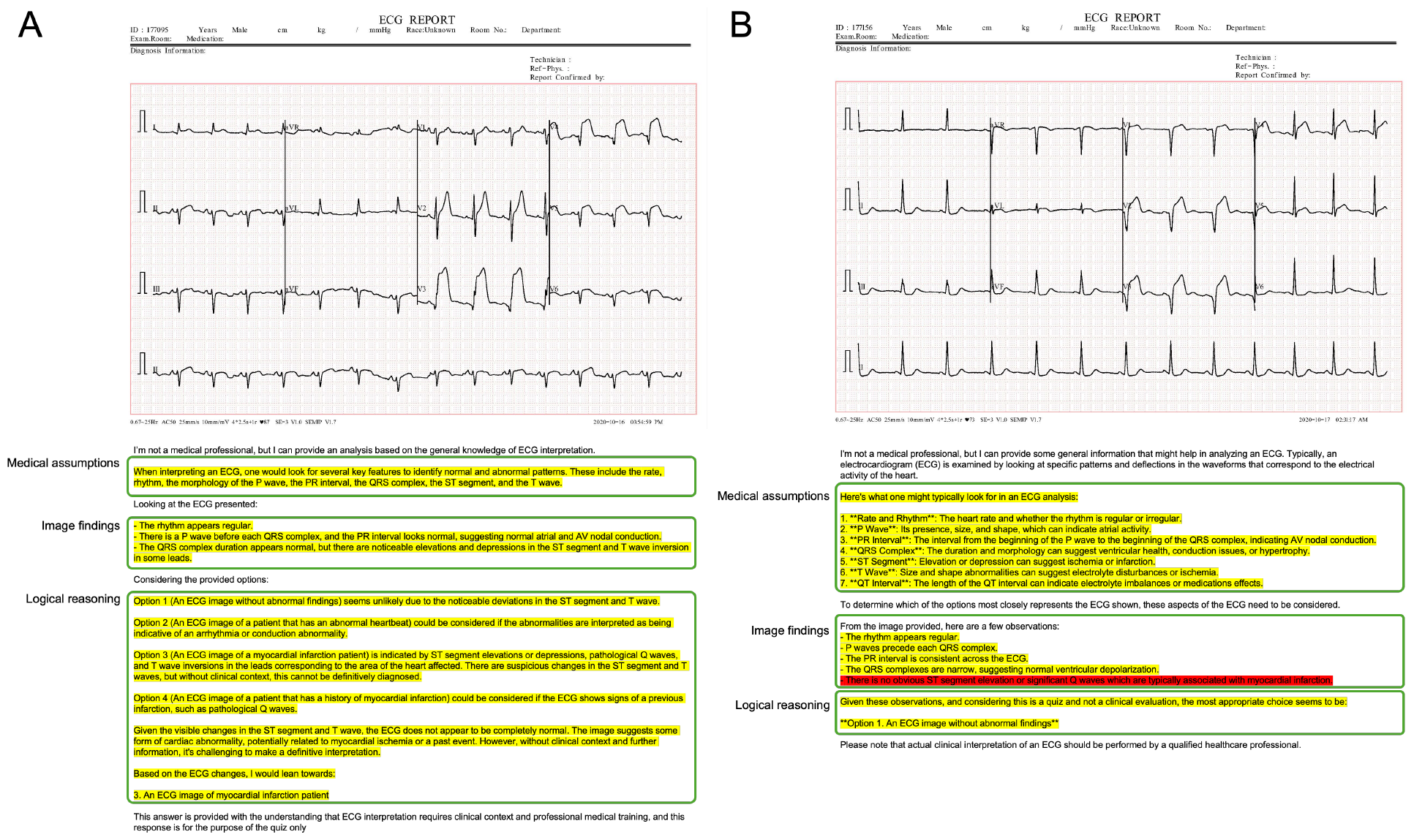
Examples of actual input images and text output by ChatGPT Plus. Both A and B are samples labeled as myocardial infarction. Yellow text indicates accurate content regarding the image, while red text indicates errors. In logical reasoning, the case of inconsistency with verbalized information was judged as abnormal, and if there was no inconsistency, there was no inconsistency in logical reasoning.

**Figure 4.**
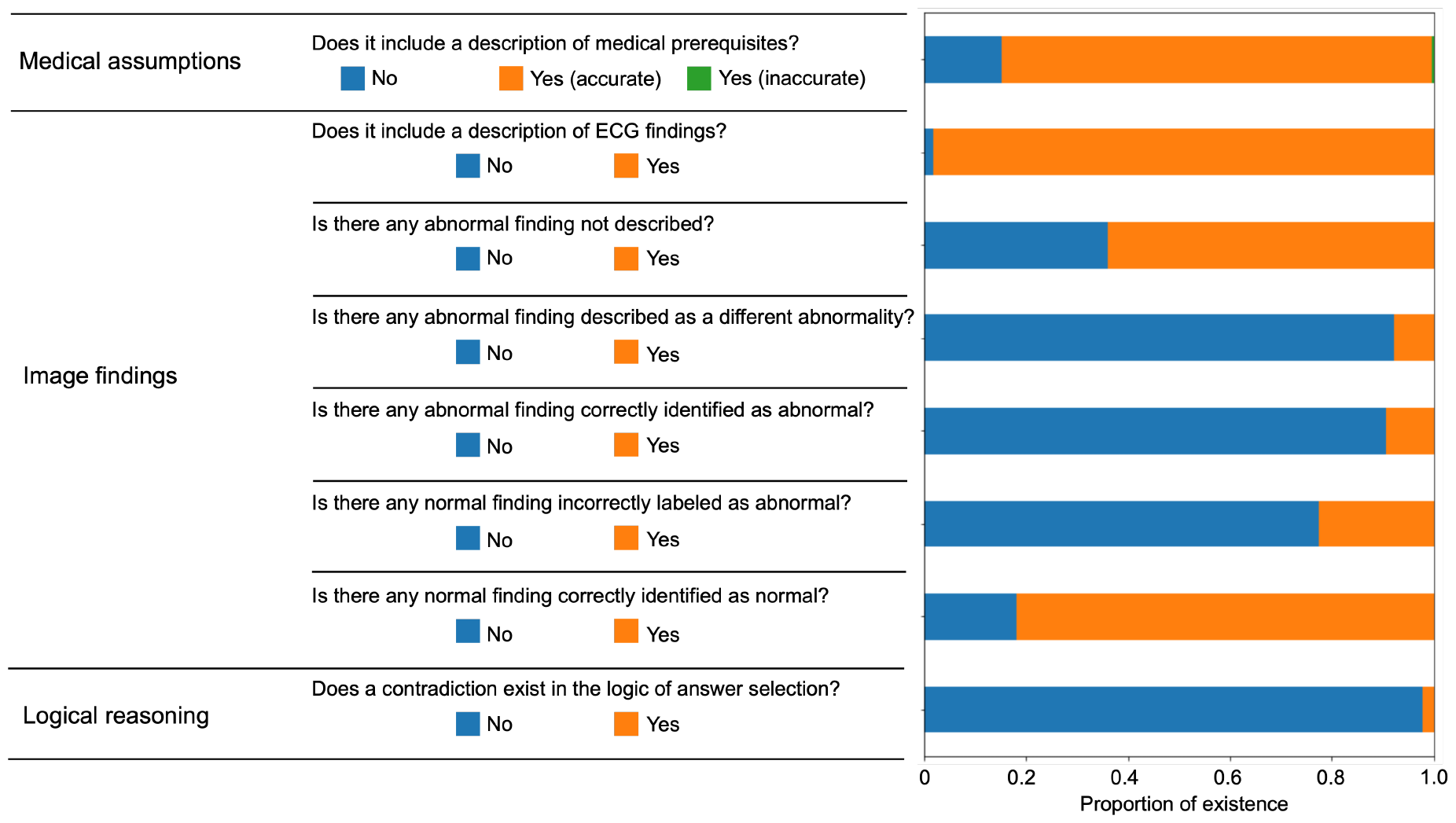
Verification results of all text outputs using ChatGPT Plus.

Figure 5 illustrates the validation outcomes of the sentences generated by ChatGPT, which are depicted individually for each label. A higher incidence of missed abnormal findings was observed in the subset of labels containing abnormalities.

**Figure 5.**
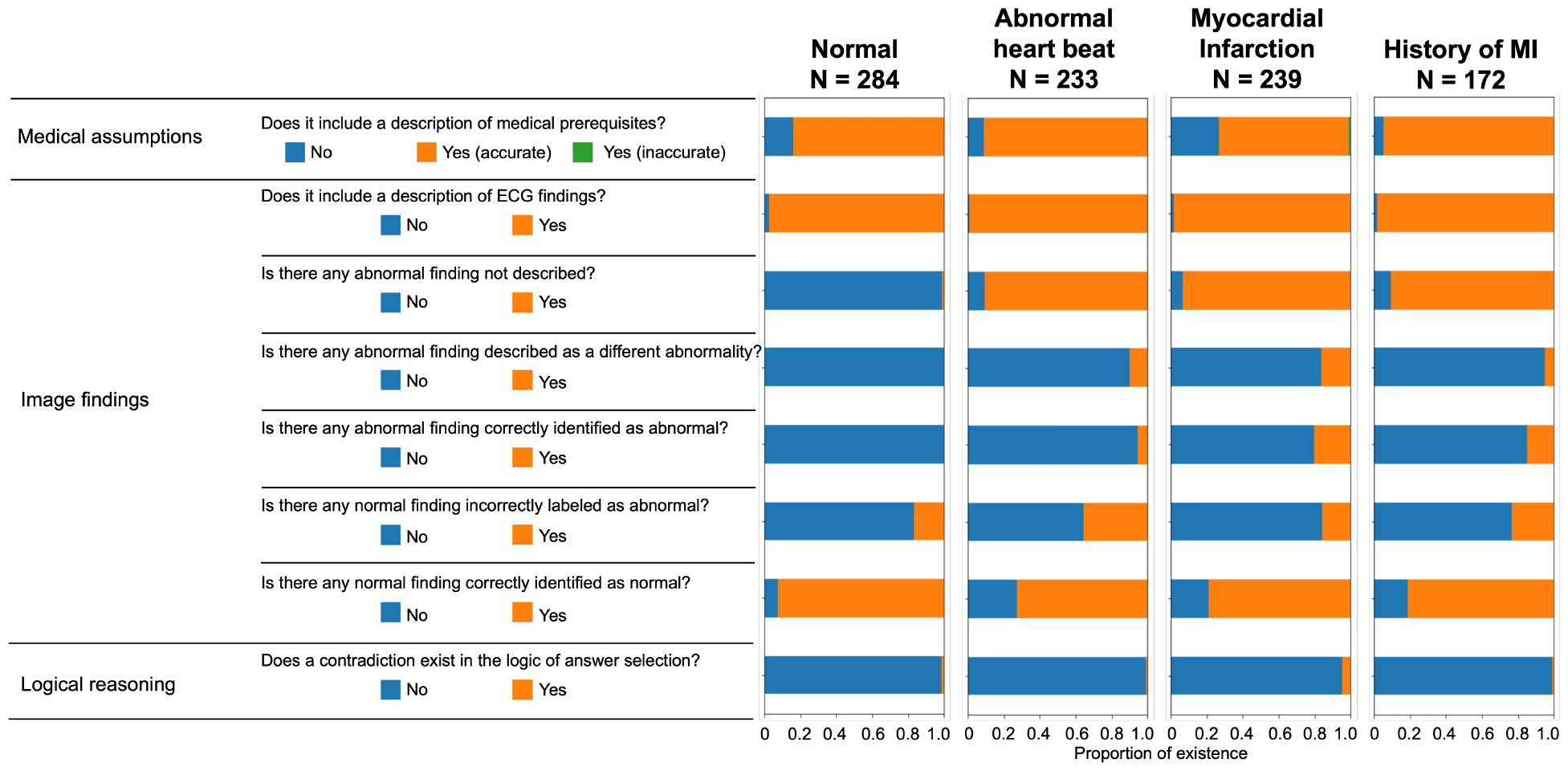
Display the validation results for each label for sentence output using ChatGPT Plus.

Figure 6 presents the validation outcomes for the sentences generated by ChatGPT Plus, categorized based on whether the correct answer choice was selected (Figure 6). Even when the correct choice was selected in the output text, a notable frequency of incorrect statements pertaining to the imaging findings remained.

**Figure 6.**
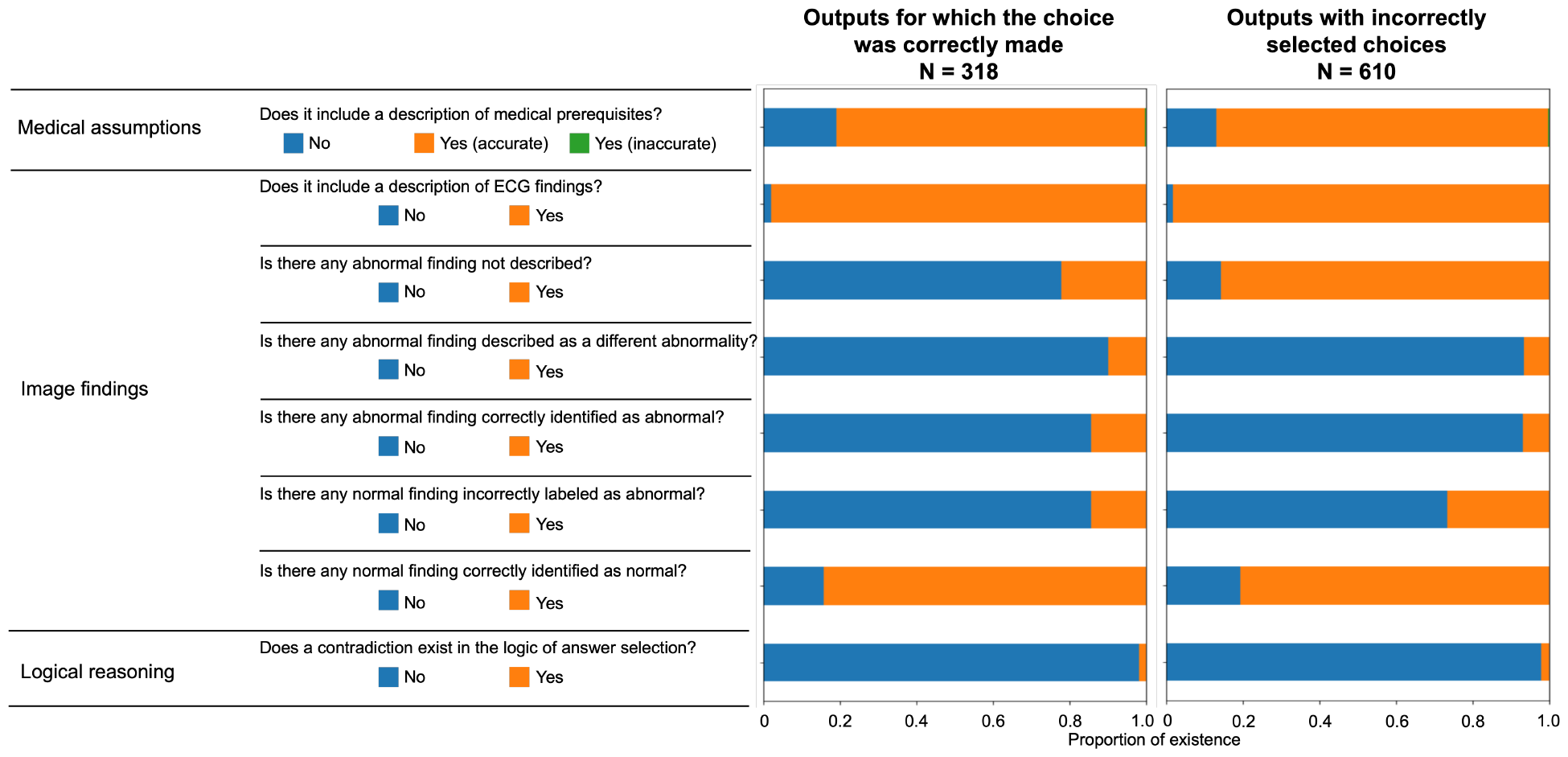
Validation results for the sentences output by ChatGPT Plus are displayed according to whether the correct answer choice was selected.

## Discussion

In this study, 12-lead ECG imaging was treated as a zero-shot VQA task and a multimodal approach for ECG interpretation was employed. The performance of all three models tested was biased in the direction of judging as normal, which was not at a practical level; however, ChatGPT Plus was slightly lower than the other two models, with a slightly higher F1 score. Additionally, a detailed validation of the ChatGPT Plus outputs revealed a higher frequency of errors in accurately extracting and verbalizing image features compared to errors in prior knowledge and logical inconsistencies in answer selection. It is hypothesized that controlling the hallucinations of input images is important for future iterations of such models. Additionally, validation of the text output by ChatGPT Plus revealed a significant number of instances in which incorrect descriptions of image features persisted despite correct answers. This underscores the importance of evaluating the ability to correctly answer visual question-answering tasks when evaluating model performance for implementation.

Hallucinations caused by LLM can be divided into factuality and faithfulness^(17,18)^. Factuality hallucinations were further divided into verifiable factual inconsistencies and fabrication. Generally, the frequency of factual inconsistency is considered the highest, and this study, in which factual inconsistency for imaging findings was the highest, is consistent with such findings. This indicates that the control of hallucinations by retrieval-augmented generation and associated methods^(20)^ may be expected in VQA of 12-lead ECGs.

One limitation of this study was that only a single dataset type was used as the input, and the number of images was limited. 12-lead ECGs are plotted in two dimensions; particularly, the sequence of leads may vary depending on the device used. Therefore, it is necessary to verify the 12-lead ECG images using different lead sequences. Additionally, ECG abnormalities are considered to be extracted for classification purposes and do not reflect the actual distribution of abnormalities. The abnormal findings in the dataset used in this study were limited, which is a limitation.

Our validation clarified the current behavior of multimodal LLMs output hallucinations in 12-lead ECG images. Currently, the accuracy of zero-shot VQA for 12-lead ECG images is still far from practical; however, it is at a stage where it is desirable to construct an appropriate evaluation method for future development.

## Data Availability

All data produced in the present study are available upon reasonable request to the authors.

https://data.mendeley.com/datasets/gwbz3fsgp8/2

